# Radiological signs supporting idiopathic intracranial hypertension in symptomatic patients with lumbar puncture opening pressure < 250 mm

**DOI:** 10.1101/2024.02.16.24302953

**Authors:** Anat Horev, Tal Eliav, Inbal Sherer, Ron Biederko, Gal Ben-Arie, Ilan Shelef, Yair Zlotnik, Tamir Regev, Erez Tsumi, Asaf Honig, Gili Givaty

**Author notes:** Equal contribution first author (Anat Horev, Tal Eliav). Equal contribution last author (Erez Tsumi, Asaf Honig). Corresponding Author: Anat Horev, MD Soroka Medical Center, Yitzhack I. Rager Blvd 151 Beer Sheva, Israel. **Conflicting interests:** The author(s) declare no potential conflicts of interest with respect to the research, authorship, and/or publication of this article.

## Abstract

**Objective:** Lumbar puncture opening pressure (LPOP) exceeding 250mmH_2_O stands as a pivotal criterion in diagnosing idiopathic intracranial hypertension (IIH), according to the revised Friedman’s criteria. However, many studies discuss the variability of LPOP, while others highlight the accuracy of radiological findings as a credible diagnostic tool for IIH. We have encountered many symptomatic patients who did not meet the LPOP criteria (with or without papilledema), despite having IIH-related symptoms and neuroimaging findings. This study aimed to investigate the radiological findings and clinical symptoms in patients suspected of having IIH without meeting the LPOP criteria.

**Methods:** We retrospectively evaluated cerebral venous sinus stenosis using the conduit Farb score (CFS) as well as other radiological findings suggestive of IIH by computed tomography venography and magnetic resonance venography in female patients ≥18 years of age with chronic headaches and suspected IIH with an LPOP <250mm.

**Results:** Our cohort comprised eighty-eight women (56 with LPOP < 200 mm H_2_O and 32 with LPOP ranging between 200-250 mmH_2_O). Among patients with LPOP in the 200-250 mmH_2_O range, 40% (12 patients) exhibited three or more radiological findings supporting IIH, compared to 17% (8 patients) in the LPOP < 200 mmH_2_O group (p=0.048).

Furthermore, cerebral venous stenosis, as measured by a Conduit Farb Score (CFS) of 5 or lower, was observed in 80% (24 patients) of the LPOP 200-250 mmH_2_O group, contrasting with 40% (19 patients) in the LPOP < 200 mmH_2_O group (p<0.001).

**Conclusion:** Cerebral venous stenosis, as well as other supportive radiological findings, were significantly more common in patients with LPOP 200-250 mmH_2_O than LPOP<200 mmH_2_O. These findings suggest that given supportive clinical and radiological evidence, patients with LPOP between 200-250 mmH_2_O, with or without papilledema, may benefit from treatment for IIH.

## Introduction

The idiopathic intracranial hypertension diagnosis criteria (IIH) were last revised and updated in 2013 by Friedman et al.(1). As stated by Friedman et al., one of the most important criteria is a lumbar puncture opening pressure (LPOP) > 250 mm H_2_0. When the LPOP is less than 250 and the other criteria are met (evidence of papilledema, normal cerebrospinal fluid (CSF) profile, normal brain parenchyma without evidence of hydrocephalus, mass, or structural lesion, and no abnormal meningeal enhancement on MRI), a diagnosis of IIH is probable but not definite.

According to the same guidelines, a diagnosis of IIH without papilledema (IIHWOP) can be made if LPOP > 250mm H_2_O, with existing unilateral or bilateral abducens palsy and three out of four positive radiological findings on magnetic resonance venography (MRV): empty sella, flattening of the posterior aspect of the globe, distention of the preoptic subarachnoid space with or without a tortuous optic nerve, transverse venous sinus stenosis. If neither papilledema nor abducens palsy is found, the diagnosis can be suggested in the presence of the three supporting radiological findings with LPOP > 250 mm. In cases where LPOP < 250mm H_2_O, the diagnosis is probable only if all other criteria (including papilledema) are met. Friedman’s criteria are considered the gold standard for diagnosis in most centers today. Since 2013, several studies on IIH have been published, many of which discuss the inaccuracy of LPOP and how it differs across age, sex, and BMI (2–4) (5–9).

Others have noted the variability of IIH and its overlap with chronic headache, estimating that a significant number of patients experience elevated intracranial pressure without papilledema(10–15). Although included in Friedman’s criteria, newer publications highlight the accuracy of radiological findings as a diagnostic tool for IIH(16–20). Morris et al. (21) pointed out the sensitivity of transverse sinus stenosis (TSS) for the diagnosis of IIH over other imaging findings. Korsbaek et al. (22) proposed that idiopathic intracranial hypertension can be diagnosed if two out of three objective findings are fulfilled: papilledema, LPOP >250 mm H_2_O, and three neuroimaging signs.

We identified a group of patients who did not meet the diagnostic criteria for IIH, most of whom did not present with papilledema, who lacked an opening pressure greater than 250 mmH_2_O. This group is often diagnosed with chronic headaches or migraines despite having IIH-related symptoms and neuroimaging findings. Some studies suggest using 200 mmH_2_O LPOP as a criterion or using prolonged manometry(9, 11, 23) to improve the underdiagnosis of patients, especially without papilledema. We aimed to investigate this specific population, comparing the radiological findings and clinical symptoms in patients with LPOP < 200 mmH_2_O to those with LPOP 200-250 mmH_2_O. We hypothesized that patients with LPOP 200-250 mmH_2_O, when compared with <200 mmH_2_O, would exhibit significant radiological similarities with IIH. This data could support the possible inclusion of patients with LPOP 200-250 mmH_2_O in the IIH criteria with additional radiological findings.

## Methods

This retrospective cohort study was approved by the Soroka University Medical Center IRB committee (18.12.2016, protocol code 0350-16). Electronic medical records of patients treated at our institution between 2003 and 2021 were obtained.

### Patient Selection

We included females ≥18 years of age with clinical, radiological, and neuro-ophthalmological suspicion for IIH admitted for completion of workup with lumbar puncture (LP). Clinical suspicion for IIH was raised in patients complaining of chronic headaches, with or without visual components (progressive visual field loss, visual obscurations or blurry vision), and tinnitus. Radiological MRV and computed tomography venography (CTV) findings suggestive of elevated ICP included scleral flattening, optic nerve sheath dilatation, slit-like ventricles, transverse-sigmoid junction stenosis. Neuro-ophthalmological suspicion for IIH consisted of new-onset papilledema.

Male patients and children under 18 were excluded as we focused on the largest IIH population: adult women of childbearing age. Patients were also excluded if they had an abnormal CSF formula on LP and/or opening CSF pressure above 250 mmH_2_O, as they met the revised Friedman’s criteria for diagnosis of IIH. We excluded patients with other pathological findings on MRV/ CTV that may cause an elevated ICP (space-occupying lesion, venous thrombosis, etc.) and patients with partial data regarding radiology, LPOP, and CSF formula.

### Data collection

CSF opening pressure, recorded headaches, papilledema, and BMI were manually collected from patients’ electronic medical records at the time of diagnosis. Other demographic information, medical history, and clinical data were automatically extracted from patient records.

The radiological data was collected by a specialized radiologist blinded to the patients’ clinical data, who reviewed their brain CTV/MRV performed closest to the time of diagnosis. The imaging was categorically graded according to the following criteria: empty sella turcica (yes/no), scleral flattening (yes/no), optic nerve ectasia over 5 mm (yes/no) and slit-like ventricles (yes/no). The imaging was also analyzed using the continuous parameters: 1) width of both optic nerves and 2) the degree of bilateral transverse sigmoid junction venous stenosis (TSS), using the conduit Farb score (CFS)^14^.

The patency grading for CFS is performed in each of the transverse sinuses (left and right) on a scale of 0 to 4, based on the degree of patency: 0 indicates 0% patency, 1 indicates less than 25% patency, 2 indicates 25-50% patency, 3 indicates 50-75% patency, and 4 indicates 75-100% patency. The scores for each side are combined to calculate the CFS (ranging from 0 to 8). A healthy, normal result is 8, indicating either no or very minimal patency. A score below 5 is considered indicative of significant stenosis.

We suggest the Idiopathic Intracranial Hypertension Radiological Scoring system (IIHRS) comprising four radiological findings: empty sella, sclera flattening, optic nerve dural ectasia, and a conduit score of 5 or less. The composite score is determined by aggregating all positive radiological findings mentioned above. The scoring system was created specifically to correlate with the revised Friedman’s criteria^1^, in which at least 3 positive radiological supporting findings are needed for IIH diagnosis in patients without papilledema.

### Statistical analysis

The statistical analysis tables present data summaries of main variables in the form of means and standard deviations for normal distributed quantitative variables and medians and interquartile range (IQR) for non-normal distributed quantitative variables (i.e., the measurements of the optic nerve sheaths), and percentages for categorical variables. A Mann-Whitney test was used to compare the FARB score between the two groups. The patient’s medical history, imaging findings such as Empty Sella and Scleral flattening, and the presence of IIH symptoms such as papilledema and tinnitus were compared between the study groups using a Pearson chi-square test.

Primary analysis of the association between the degree of sinus stenosis (measured in Farb scale) and cerebrospinal fluid (CSF) opening pressure was constructed using multivariate Poisson regressions with opening pressure as the dependent variable. Statistical significance was set at a two-sided p<0.05 or confidence interval of 95%. Reported p-values were rounded to three decimals. Statistical analysis was performed using R-studio version 4.1.1 (R-foundation).

## Results

A total of 88 female patients were included: 20 patients with normal BMI (22.4%), 28 patients with BMI between 25-30 (31.4%), and 41 patients with BMI over 30 (46%). Thirty-two patients with a mean age of 33.3 (10.4) had LPOP 200-250 mmH_2_O, and 56 patients with a mean age 35.6 (13) had LPOP< 200 mmH_2_O. No differences were found between the two groups in terms of age at hospitalization, BMI, smoking status, oral contraceptive use, as well as medical history (Table 1). All patients were women of child bearing age who came to medical attention because of chronic headaches. No significant difference was found between groups in other IIH-related symptoms, such as visual complaints and tinnitus (Table 1).

**Table 1.** Patient characteristics.

|  | < 200 mmH<sub>2</sub>O<br>(N=56) | 200-250<br>mmH<sub>2</sub>O<br>(N=32) | p-value |
| --- | --- | --- | --- |
| **Age at admission, years** |  |  |  |
| Mean (SD) | 35.6 (13.0) | 33.3 (10.4) | 0.521 |
| Median [Min, Max] | 35.0 [18.0, 63.0] | 33.0 [18.0, 66.0] |  |
| **BMI, kg/m2** |  |  |  |
| Mean (SD) | 29.4 (5.63) | 30.0 (5.75) | 0.627 |
| Median [Min, Max] | 28.9 [14.6, 43.0] | 30.7 [19.9, 43.4] |  |
| **Smoking status n (%)** |  |  | 0.291 |
| No | 14 (25.0%) | 7 (21.9%) |  |
| Yes | 13 (23.2%) | 8 (25.0%) |  |
| In the past | 1 (1.8%) | 3 (9.4%) |  |
| **Oral contraceptive pills n (%)** | 12 (21.4%) | 6 (18.8%) | 0.895 |
| **Psychiatric history n (%)** |  |  |  |
| Anxiety | 21 (37.5%) | 10 (31.3%) | 0.72 |
| Depression | 14 (25.0%) | 4 (12.5%) | 0.261 |
| Schizophrenia | 2 (3.6%) | 0 (0%) | 0.735 |
| Other psychiatric diagnosis | 12 (21.4%) | 2 (6.3%) | 0.116 |
| **Medical history n (%)** |  |  |  |
| Congestive heart failure | 1 (1.8%) | 0 (0%) | 1 |
| Peripheral vascular disease | 1 (1.8%) | 0 (0%) | 1 |
| Cerebrovascular disease | 6 (10.7%) | 3 (9.4%) | 1 |
| COPD | 8 (14.3%) | 3 (9.4%) | 0.738 |
| Peptic ulcer disease | 3 (5.4%) | 0 (0%) | 0.471 |
| Mild liver disease | 3 (5.4%) | 2 (6.3%) | 1 |
| Diabetes | 6 (10.7%) | 3 (9.4%) | 1 |
| Hypertension | 9 (16.1%) | 4 (12.5%) | 0.887 |
| **Papilledema at hospitalization n (%)** | 5(8.9%) | 6 (18.8%) | 0.407 |
| **Tinnitus at hospitalization n (%)** | 9 (16.1%) | 5 (15.6%) | 1 |
| **Visual complaints n (%)** | 29 (51.8%) | 22 (68.8%) | 0.215 |
| **Visual complaints n (%)** |  |  | 0.063 |
| Visual obscurations | 23 (41.1%) | 16 (50.0%) |  |
| Blurred vision, nonspecific | 2 (3.6%) | 0 (0%) |  |
| Other nonspecific visual complaints | 4(6.2%) | 6 (18.8%) |  |

\* p <.05

Most patients lacked papilledema, with no significant differences found between groups (62.5% LPOP 200-250 mmH_2_O and 69.9% LPOP<200mmH_2_O, p=0.407). Neuroradiological findings are presented in Table 2. Among patients with LPOP 200-250mmH_2_O, 62.5%% exhibited an empty sella compared with 33.9% of those with LPOP<200 mmH_2_O (p = 0.018). The CFS was significantly lower in the LPOP group of 200-250mmH_2_O (mean=3.73, SD=1.98) versus the LPOP <200 mmH_2_O group (mean=5.72, SD=2.35, p-value < 0.001).

**Table 2.** Neuroradiological findings.

|  | LPOP< 200<br>mmH<sub>2</sub>O<br>(N=56) | LPOP: 200-250 mmH<sub>2</sub>O<br>(N=32) | p-value |
| --- | --- | --- | --- |
| **Empty sella** n (%) | 19 (33.9%) | 20 (62.5%) | 0.018* |
| **Slit-like vent** n (%) | 9 (16.1%) | 9 (28.1%) | 0.283 |
| **Scleral flattening** n (%) | 5 (8.9%) | 7 (21.9%) | 0.179 |
| **Optic nerve dural ectasia** n (%) | 18 (32.1%) | 17 (53.1%) | 0.1 |
| **Optic nerve width, right millimeters** |  |  |  |
| Mean (SD) | 6.28 (0.752) | 6.56 (0.629) | 0.234 |
| Median [Min, Max] | 6.00 [5.00, 8.00] | 6.50 [6.00, 8.00] |  |
| **Optic nerve width, left millimeters** |  |  |  |
| Mean (SD) | 6.50 (0.857) | 6.88 (0.619) | 0.109 |
| Median [Min, Max] | 6.00 [5.00, 8.00] | 7.00 [6.00, 8.00] |  |
| **Farb score, right** |  |  |  |
| Mean (SD) | 2.96 (1.32) | 1.77 (1.14) | <0.001<br>* |
| Median [Min, Max] | 4.00 [0, 4.00] | 1.50 [0, 4.00] |  |
| **Farb score, left** |  |  |  |
| Mean (SD) | 2.77 (1.37) | 1.97 (1.35) | 0.013* |
| Median [Min, Max] | 3.00 [0, 4.00] | 2.00 [0, 4.00] |  |
| **Conduit Farb score (CFS)** |  |  |  |
| Mean (SD) | 5.72 (2.35) | 3.73 (1.98) | <0.001<br>* |
| **CFS under 5 (significant venous stenosis)** n (%) | 19 (40.4%) | 24 (80%) | 0.001* |
| **IIHRS** |  |  |  |
| Mean (SD) | 1.09 (1.13) | 2.13 (1.29) | <0.001<br>* |
| Median [Min, Max] | 1.00 [0, 4.00] | 2.00 [0, 4.00] |  |
| **IIHRS above 3** n (%) | 8 (14.3%) | 12 (37.5%) | 0.025 |

\* p < .05

Venous sinus stenosis calculated by CFS (<5) was found in 80% of those with LPOP 200-250mmH_2_O and 40.4%% of LPOP<200 mmH_2_O (p<0.001). IIHRS was found to be significantly higher in the LPOP 200-250mm H_2_O group (p<0.001). IIHRS score above 3 was found in 37.5% of the LPOP 200-250mm H_2_O group versus 14.3% in the <200 mm H_2_O group (p=0.025).

The results of multiple Poisson regressions are presented in Table 3. We observed a negative association between CFS and CSF opening pressure (IRR = 0.75; [95% confidence interval, CI, 0.59-0.94]; p=0.01). Another negative association was found between the CFS and scleral flattening (IRR = 0.59; [95% confidence interval, CI, 0.38-0.88]; p=0.01). A negative relationship was found between the CFS and the empty sella, as well, however, it was not statistically significant (IRR = 0.82; [95% confidence interval, CI, 0.65-1.03]; p=0.09). The association between the CFS and optic nerve dural ectasia showed no difference (IRR = 1.00; [95% confidence interval, CI, 0.78-1.26]; p=0.9).

**Table 3.**
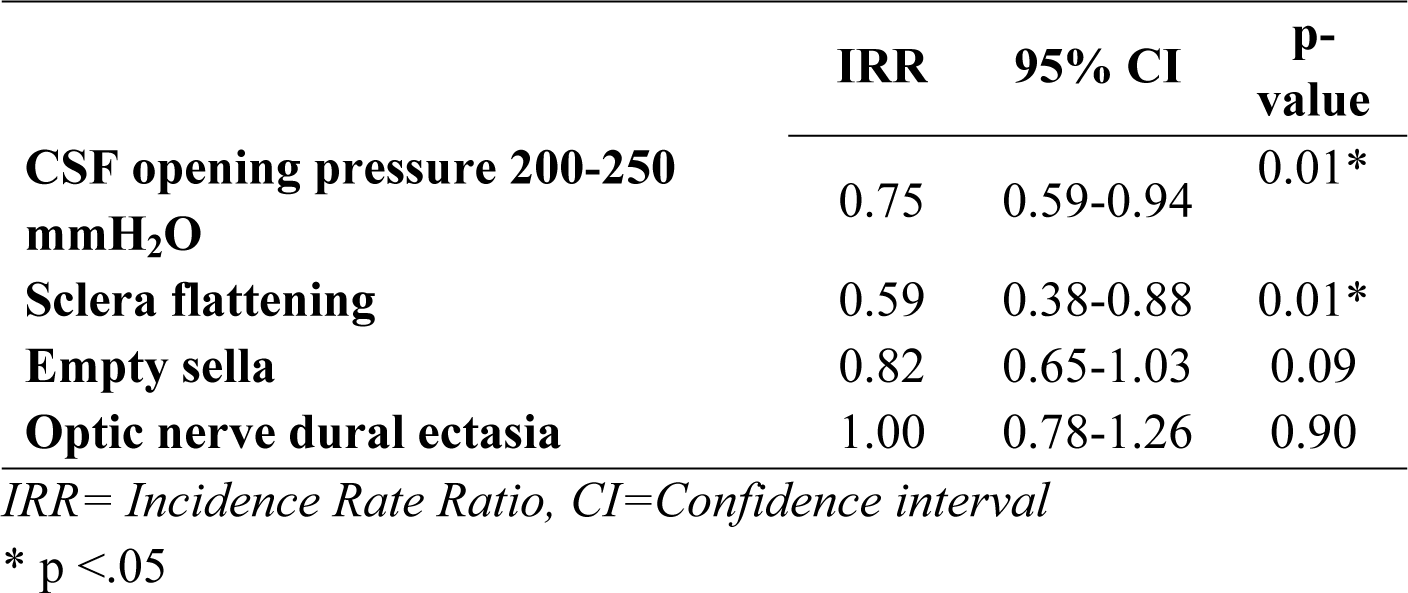
Poisson regression model: association between CFS score and patient presentation.

|  | IRR | 95% CI | P-value |
| --- | --- | --- | --- |
| **CSF opening pressure 200-250 mmH<sub>2</sub>O** | 0.75 | 0.59-0.94 | 0.01* |
| **Sclera flattening** | 0.59 | 0.38-0.88 | 0.01* |
| **Empty sella** | 0.82 | 0.65-1.03 | 0.09 |
| **Optic nerve dural ectasia** | 1.00 | 0.78-1.26 | 0.90 |

*IRR= Incidence Rate Ratio, CI=Confidence interval*
\* $p < .05$

## Discussion

In this retrospective study, we analyzed chronic headache patients with suspected IIH who did not meet Friedman’s LPOP criteria >250 mmH_2_O. All 88 participants were women, 56 of whom had an LPOP< 200 mmH_2_O and 32 who had LPOP 200-250 mmH_2_O. Seventy-seven (87.5%) patients did not have papilledema on presentation. When comparing between groups, we found that patients with LPOP 200-250 mmH_2_O were significantly more likely to have an empty sella than those <200mmH_2_O (62.5% versus 33.9%, p=0.018). Our results further demonstrated that 80% of LPOP 200-250 mmH_2_O had CFS values supporting severe TSS <5 (mean 3.73). In contrast, of those with opening pressure <200 mmH_2_O, only 40.4% demonstrated severe TSS (<0.001).

Farb et al.(17) first described the correlation between venous stenosis and IIH in 2003. He found that 93% of patients with IIH had a combined conduit score less than 5, indicating severe stenosis. Morris et al.(21) compared TSS in IIH patients to non-IIH patients. He found that 83% of the IIH patients had bilateral TSS versus 4% of the non-IIH patients and 53% of the IIH patients had empty sella on MRI versus 5% in non-IIH patients. Furthermore, 63% of the IIH patients had optic nerve dilatation compared with 4% of the non-IIH patients.

When comparing our study group with LPOP 200-250 mmH_2_O to Morris et al.’s confirmed IIH patients, we show similar radiological findings (76% with severe VSS, 62.5% with empty sella and 53.1% with optic nerve dural ectasia). This is in accordance with existing literature that describes a similar percentage of IIH patients with these imaging results(16, 23–25), supporting the similarity between patients with 200-250 and >250 LPOP.

Although those with LPOP 200-250mmH_2_O (largely IIHWOP) carry very suggestive clinical and radiological findings of IIH, they do not meet Friedman’s criteria.(^1^) According to the Friedman et al., in patients without papilledema, IIH diagnosis is possible if all other criteria are met: LPOP>250 mmH_2_O and abducens palsy or three out of four positive radiological supportive findings: empty sella turcica, scleral flattening, optic nerve dural ectasia and transverse venous stenosis. In our study we showed that 40% of the LPOP 200-250 mmH_2_O group had at least three positive radiological findings to support IIH (IIHRS>3). While we did not find abducens palsy in our patients, it may have been overlooked in some patients, although it is unclear to what extent.

Cerebral venous stenosis is the most sensitive sign for IIH(21, 26{Quan, 2019 #350)(21, 26, 27) and 80% of patients with LPOP 200-250 mmH_2_O in our study had CFS<5, providing further support that this group, or at least a part of it, should be included as IIH patients.

Prior to the publication of Friedman’s revised criteria in 2013, the normal upper limit for LPOP was defined as 200mmH_2_O by the International Headache Society(^28^). Three studies addressed normative values for CSF opening pressure and found average CSF pressure values of 155, 170, and 193 mmH_2_O(29-31).

Norager et al.(32) published a review of the references for LPOP based on six studies, none of which showed values greater than 200 mm(28). However, evidence has also been published that supports 250 mmH_2_O as the normal upper limit(29, 30, 33, 34). Many studies attempted to resolve this uncertainty by discussing fluctuations affecting LPOP(23, 35).

Others have suggested that opening pressure is affected by body position, age, BMI, anesthesia, needle size, and measurement technique(3–7, 31, 36, 37). According to Wang et al. 12.5% of patients showed an increase of more than 50mm on LPs repeated within 2.5 years(31). This was also seen in a study by Kilgore et al. who showed significant differences in LPOP on repeated tests(8). Together, these studies suggest a high range of variability and inaccuracy of a test that is both uncomfortable for the patient and carries a significant risk of complication(32, 38).

Bono et al. showed that an hour-long CSF manometry could be considerably more informative than recording a single spot opening pressure. They further showed that, especially in patients with bilateral venous stenosis, the LPOP may be less than 250 mmH_2_O but can reach higher pressures if monitored for 60 minutes(23). A review of 27 LPOP studies found that it was highly variable amongst patients and concluded that using a single value of ICP to distinguish normal values from pathological ones may not be biologically natural(32).

Our study highlights a significant patient population with LPOP between 200-250 mmH_2_O who have radiological and clinical findings supporting the diagnosis of IIH, most of whom had no papilledema at presentation. This population is not diagnosed with IIH and is therefore not treated accordingly. It is important to note that in the group of patients with LPOP <200H_2_O, a non-negligible number of patients had supportive radiological findings for IIH (although found to be significantly less common).

Multiple studies support the accuracy of radiological findings in the diagnosis of IIH. The most accurate findings were found to be VSS, empty sella, and scleral flattening(19, 20, 22, 26, 39, 40). It was also published that up to 48% of chronic headache sufferers have bilateral cerebral venous stenosis(41, 42). Is very likely that our cohort supports the underdiagnosis of IIH in patients with chronic headaches and cerebral venous stenosis that did not undergo lumbar punctures. This population is likely to have elevated intracranial pressure that is undiagnosed and, therefore, undertreated. Recent data suggests that it is advantageous to consider IIH on a spectrum(13, 43, 44). This is backed by the importance of the radiological findings and uncertainties surrounding the accuracy of the LPOP. Consequently, we, alongside many other researchers, propose revising the diagnostic criteria for IIH.

We present the first recent study directly comparing patients with suspected IIH and an LPOP of 200-250 mmH_2_O to those with pressures <200 mmH_2_O. While this group has been mentioned in some studies, it has not been their primary focus. Although retrospective and based on a small cohort, our research brings attention to a subpopulation that may be experiencing IIH. This specific population may be underdiagnosed and, therefore, may benefit from treatment of IIH. Further large-scale studies are needed to support our conclusions.

## Statement of authorship

Conception and design: Horev, Eliav, Honig Drafting of manuscript: Horev, Eliav, Sherer, Givaty Data collection and analysis: Biederko, Ben-Arie, Shelef, Regev, Tsumi Critical revision: Horev, Eliav, Honig, Zlotnik All authors approved the final manuscript.

## Data Availability

All relevant data are within the manuscript and its Supporting Information files.

